# Homologous recombination deficiency in ovarian high-grade serous carcinoma by self-reported race

**DOI:** 10.1101/2025.01.21.25320918

**Authors:** Katherine A. Lawson-Michod, Courtney E. Johnson, Mollie E. Barnard, Natalie Davidson, Lindsay J. Collin, David A. Nix, Chad Huff, Andy Berchuck, Lucas A. Salas, Casey Greene, Jeffrey R. Marks, Lauren C. Peres, Jennifer A. Doherty, Joellen M. Schildkraut

## Abstract

**Background:** Approximately half of ovarian high-grade serous carcinomas (HGSC) have homologous recombination deficiency (HRD). However, HRD is not well-characterized in Black individuals.

**Objective:** To characterize HGSC HRD by self-reported race and evaluate whether differences in HRD are associated with ovarian cancer mortality.

**Study population:** Cohort study using data collected from two population-based case-control studies of ovarian cancer. Cases were selected based on self-reported race (178 Black, 123 White) and pathologically-confirmed HGSC.

**Exposures:** HRD features identified using matched tumor-normal whole-exome DNA sequencing and categorized as germline or somatic variants in homologous recombination pathway genes, or the SBS3 HRD-associated signature.

**Outcomes:** Median difference and 95% confidence intervals (CI) for age at diagnosis and tumor mutation burden, and age and stage-adjusted hazard ratios (HR) and 95%CIs for survival, comparing individuals with an HRD feature to those without, separately by self-reported race.

**Results:** More of the germline and somatic variants detected among Black individuals compared with White individuals were unannotated or variants of uncertain significance (VUS; germline 65% versus 45%; somatic 62% versus 50%, respectively). While the prevalences of many HRD features were similar between Black individuals and White individuals, Black individuals had a higher prevalence of the HRD signature identified using de novo mutational signature analysis (40% versus 29%) and germline *BRCA2* variants (8% versus 2%) compared with White individuals. We observed that among Black individuals, *BRCA2* variants were associated with better survival (somatic HR=0.23, 95%CI 0.07–0.76; germline HR=0.48, 95%CI 0.22–1.03), while germline *BRCA1* variants were associated with worse survival (HR=2.11, 95%CI 1.14–3.88). When we restricted to VUS and unannotated variants, we observed similar associations with survival for *BRCA*2 among Black individuals (somatic HR=0.18, 95%CI 0.04-0.75; germline HR=0.40, 95%CI 0.15–1.09).

**Conclusions and Relevance:** HRD testing informs precision-based medicine approaches that improve outcomes, but a higher proportion of VUS among Black individuals may complicate referral for such care. Our findings emphasize the importance of recruiting diverse individuals in genomics research and better characterizing VUS.

## INTRODUCTION

Ovarian cancer is the sixth leading cause of cancer-related death in women and is comprised of multiple histotypes.^1–3^ High-grade serous carcinoma (HGSC)—the most common histotype—is usually diagnosed at a late stage. While HGSC initially responds to surgical debulking and chemotherapy, most patients relapse, resulting in poor survival.^4^ Approximately 50% of HGSC have genetic or epigenetic alterations in homologous recombination pathway genes, most commonly in *BRCA1* and *BRCA2*, leading to homologous recombination deficiency (HRD).^5^ HRD is associated with higher tumor mutation burden (TMB) and these tumors are more responsive to chemotherapy leading to better patient survival.^6–9^ Genetic testing to inform risk-reducing surgery has contributed to prevention of ovarian cancer among *BRCA* carriers and tumor HRD testing has informed the use of poly(ADP-ribose) polymerase (PARP) inhibitors which have shown effectiveness in improving progression-free survival in patients with HRD-positive ovarian cancer.^10,11^

The survival benefit from these advances may not be equally distributed across all racial and ethnic groups. While ovarian cancer mortality is declining, the rate of decrease among Black individuals is half that of White individuals.^12^ Compared with White individuals, Black individuals are less likely to be referred for genetic testing due to family history, less likely to complete testing following a referral, and less likely to undergo prophylactic salpingo-oophorectomy if they have a pathogenic risk variant.^13–18^ Further, our understanding of HRD in HGSC is informed by research in predominately White populations and therefore enriched for HRD features specific to White populations.^19–26^ This may contribute to higher rates of variants of uncertain significance (VUS) among Black individuals, further complicating treatment referral as VUS cannot be used to inform clinical decision-making.^23,24,27–29^

Characterizing HRD among Black individuals with HGSC is of particular importance given prior studies that report HRD may be more prevalent in tumors from Black individuals with various cancer types.^23,24,30–32^ While prior studies observed variation in germline *BRCA1* and *BRCA2* variants by geography and ancestry, characterization of this variation is still limited and few studies have evaluated population differences in somatic mutations, non-*BRCA* genes, or within specific ovarian cancer histotypes.^19–26^ Here, we characterize HRD in 178 Black and 123 White individuals with HGSC and evaluate the degree to which HRD features are associated with tumor mutation burden, age at diagnosis, and survival in each population.

## METHODS

### Study Population

Our cohort included individuals with pathologically confirmed HGSC, 178 who self-reported Black race (“SchildkrautB”) and 123 who self-reported White race (“SchildkrautW”), enrolled in one of two population-based case-control studies of newly diagnosed epithelial ovarian cancer, the African American Cancer Epidemiology Study (AACES), and the North Carolina Ovarian Cancer Study (NCOCS).^33–36^ AACES included individuals ages 20 to 79 years, self-reporting Black race, and diagnosed between 2010–2015. Individuals eligible for NCOCS were ages 20 to 74 years at diagnosis and diagnosed between 1999–2005. Institutional Review Board (IRB) approval was obtained for patient recruitment, sample collection, and research. Written informed consent was received from all NCOCS participants. All AACES participants provided verbal consent for interview and written consent for blood or saliva sample collection. Both study populations signed release forms for medical records and tissue samples. HGSC histotype was confirmed by centralized pathology review. Age at diagnosis was extracted from questionnaires and pathology reports. Stage, debulking status, and receipt of neoadjuvant therapy were obtained from medical records, pathology reports, and cancer registries. Vital status was determined from cancer registries, direct physician contact, and the National Death Index. First-degree family history of breast and ovarian cancer was self-reported in questionnaires. White individuals were matched to Black individuals on 5-year age categories and oversampled for early stage prior to tumor sequencing (eFigure 1).

**Figure 1.**
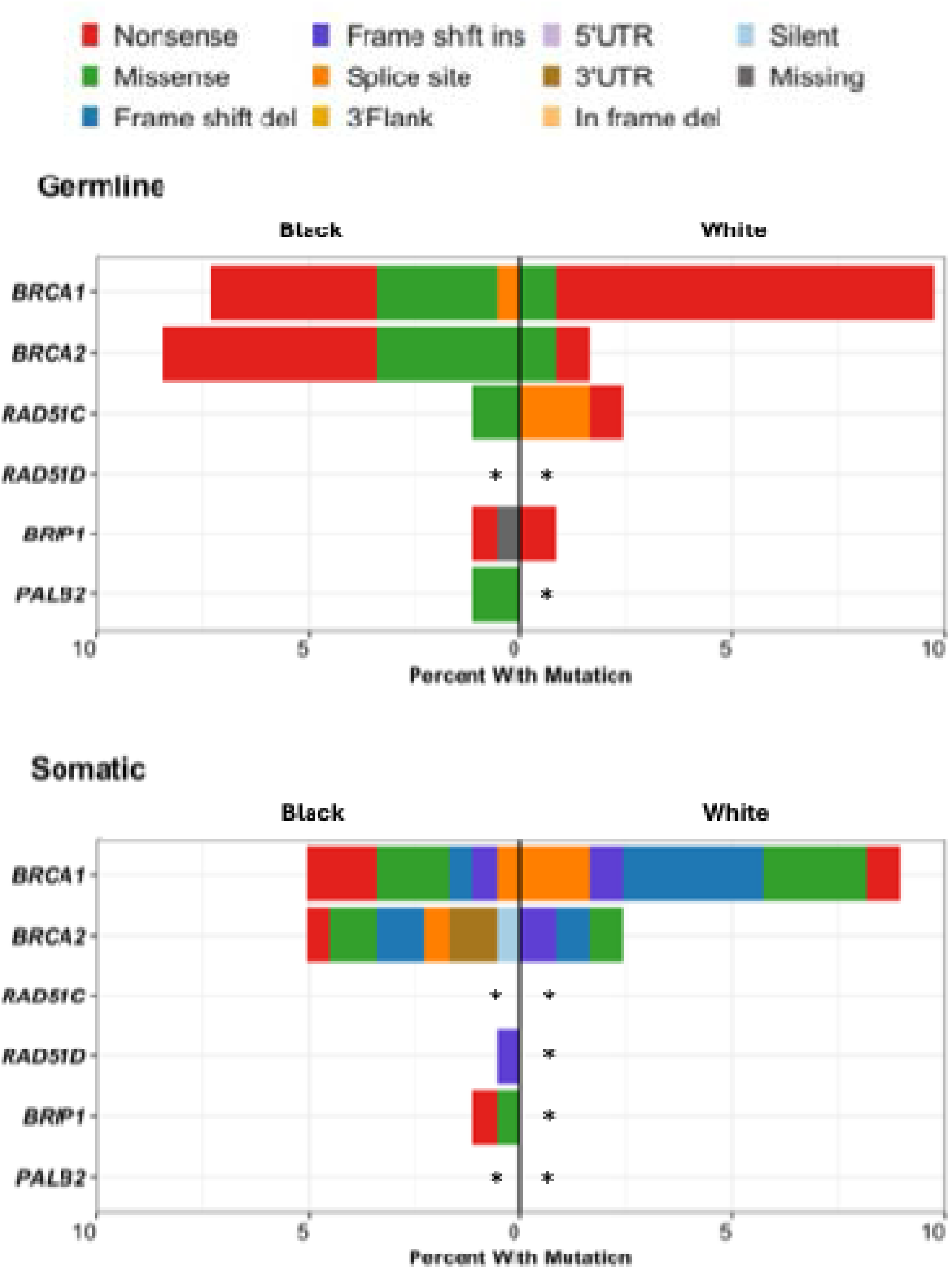
Germline variants and somatic mutations in homologous recombination pathway genes prioritized in the 2020 American Society of Clinical Oncology (ASCO) recommendation for genetic testing in ovarian cancer. A) Percentages of germline variants (top panel) or somatic mutations (bottom panel) and the variant classifications in Black individuals (left) and White individuals (right). Genes with * had no identified variants. Abbreviation: UTR, untranslated region

### HRD Features

Germline and somatic variants were identified through whole-exome sequencing in matched germline and formalin-fixed paraffin embedded tumor samples. We excluded benign and likely benign variants and restricted to germline variants with a quality score of ≥99.9 for SNVs and 98.0 for INDELs, read depth and genotype quality ≥10, alternative read proportion between 20%–80%, and minor allele frequency <0.5% and somatic variants passing Strelka’s empirical variant score model with tumor read depth ≥30, tumor allele fraction ≥15%, normal allele frequency <5%, quality score ≥15, and population frequency <1%.

In our primary analyses we included genes involved in the homologous recombination pathway and recommended by the 2020 American Society for Clinical Oncology (ASCO) guidelines to be prioritized in epithelial ovarian cancer germline genetic testing: *BRCA1, BRCA2, RAD51C, RAD51D, BRIP1, PALB2*.^37^ We categorized HRD features as germline variants or somatic mutations in (1) *BRCA1*; (2) *BRCA2*; (3) any other prioritized gene in the ASCO guidelines (labeled as *any non-BRCA ASCO prioritized gene* in tables and figures) (4) any ASCO prioritized gene (*any ASCO prioritized gene*); (5) *HRD signature* identified using *de novo* mutational signature analysis using SigProfilerExtractor; (6) *any HRD* which included individuals with one or more of the features described above.^35,38^ We reported the prevalence of potentially pathogenic variants in these categories and the proportion of potentially pathogenic variants annotated as pathogenic/likely pathogenic in ClinVar and VUS or unannotated in ClinVar separately by self-reported race.

Because the ASCO prioritized gene list is based on evidence for susceptibility not outcomes, we performed sensitivity analyses including additional homologous recombination pathway genes: *ATM*, *ATR*, *ATRX, BARD1, BLM, CHEK1, CHEK2, FANCC, FANCD2, FANCE, FANCF, FANCG, FANCI, FANCL, FANCM, MRE11A, NBN, PALB2, RAD50, RAD51, RAD51B, RAD52, RAD54L, RPA1*.^37^

### Statistical Analysis

We estimated the difference in the median tumor mutation burden (TMB) defined by the number of somatic mutations per megabase (Mb) and age at diagnosis for individuals with an HRD feature compared with those without using unadjusted linear models and calculating 95%CI, separately by self-reported race. We used Cox proportional hazards regression to calculate HRs for associations between the presence of the HRD feature and all-cause mortality adjusting for age and stage, separately by self-reported race. We tested for an interaction with race on the multiplicative scale. We repeated analyses restricting to VUS and unannotated variants. All analyses were performed in R version 4.2.2 (Vienna, Austria).

## RESULTS

### Study Population

Debulking status was available for 119 Black individuals and 34 White individuals, and among those, optimal debulking was achieved in 61% and 88%, respectively. A first-degree family history of ovarian cancer was reported by 9% of individuals in both groups while a first-degree family history of breast cancer was reported by 34% of Black individuals and 20% of White individuals (Table 1).

**Table 1.** Characteristics of high-grade serous ovarian cancer cohort, by self-reported race.

| Participant characteristics | Black<br>(n=178) |  | White<br>(n=123) |  | p-value<sup>□</sup> |
| --- | --- | --- | --- | --- | --- |
|  | No. | (%) | No. | (%) |  |
| **Age at Diagnosis<sup>†</sup>** |  |  |  |  | 0.93 |
| 35–39 | 5 | (2.8%) | 3 | (2.4%) |  |
| 40–44 | 12 | (6.7%) | 8 | (6.5%) |  |
| 45–49 | 17 | (9.6%) | 16 | (13%) |  |
| 50–54 | 28 | (16%) | 18 | (15%) |  |
| 55–59 | 37 | (21%) | 24 | (20%) |  |
| 60–64 | 31 | (17%) | 26 | (21%) |  |
| 65–69 | 20 | (11%) | 14 | (11%) |  |
| ≥ 70 | 28 | (16%) | 14 | (11%) |  |
| **Stage<sup>†</sup>** |  |  |  |  | 0.13 |
| FIGO I/II | 35 | (20%) | 16 | (13%) |  |
| FIGO III/IV | 141 | (80%) | 105 | (87%) |  |
| Unknown | 2 |  | 2 |  |  |
| **Debulking Status** |  |  |  |  | 0.003 |
| Optimal | 73 | (61%) | 30 | (88%) |  |
| Suboptimal | 46 | (39%) | 4 | (12%) |  |
| Unknown | 59 |  | 89 |  |  |
| **First-Degree Family History** |  |  |  |  |  |
| Ovarian Cancer | 14 | (8.8%) | 10 | (8.6%) | 0.97 |
| Breast Cancer | 55 | (34%) | 23 | (20%) | 0.01 |
| **Germline<sup>‡</sup>** |  |  |  |  |  |
| *BRCA1* | 13 | (7.3%) | 12 | (9.8%) | 0.45 |
| *BRCA2* | 15 | (8.4%) | 2 | (1.6%) | 0.012 |
| *RAD51C* | 2 | (1.1%) | 3 | (2.4%) | 0.4 |
| *RAD51D* | 0 | (0%) | 0 | (0%) |  |
| *BRIP1* | 2 | (1.1%) | 1 | (0.8%) | 1 |
| *PALB2* | 2 | (1.1%) | 0 | (0%) | 0.52 |
| Any Non-*BRCA* ASCO Prioritized | 6 | (3.4%) | 4 | (3.3%) | 1 |
| Gene<sup>§</sup> |  |  |  |  |  |
| Any ASCO Prioritized Gene | 33 | (19%) | 18 | (15%) | 0.37 |
| **Somatic<sup>‡</sup>** |  |  |  |  |  |
| *BRCA1* | 9 | (5.1%) | 11 | (8.9%) | 0.18 |
| *BRCA2* | 8 | (4.5%) | 3 | (2.4%) | 0.53 |
| *RAD51C* | 0 | (0%) | 0 | (0%) |  |
| *RAD51D* | 1 | (0.6%) | 0 | (0%) | 1 |
| *BRIP1* | 2 | (1.1%) | 0 | (0%) | 0.52 |
| *PALB2* | 0 | (0%) | 0 | (0%) |  |
| Any Non-*BRCA* ASCO Prioritized | 3 | (1.7%) | 0 | (0%) | 1 |
| Gene<sup>§</sup> |  |  |  |  |  |
| Any ASCO Prioritized Gene | 19 | (11%) | 14 | (11%) | 0.85 |
| **HRD Signature** | 72 | (40%) | 36 | (29%) | 0.047 |
| **Any HRD** | 92 | (52%) | 58 | (47%) | 0.44 |
| **Median TMB (Range)** | 1.38 (0.22, 4.42) |  | 1.32 (0.08, 4.52) |  | 0.11 |

Abbreviation: FIGO, International Federation of Gynecology and Obstetrics; HRD, homologous recombination deficiency; TMB, tumor mutation burden; ASCO, American Society of Clinical Oncology; PV, pathogenic variant; PPV, potentially pathogenic variant; PPGV, potentially pathogenic germline variant.
<sup>†</sup>Unless otherwise indicated.
<sup>‡</sup>Frequency matched on 5-year age category and White cases oversampled for early stage prior to sequencing.
<sup>§</sup>HRD categories are not mutually exclusive. Some cases had variants in >1 genes.
<sup>§</sup>Non-*BRCA* category includes alterations in *RAD51C*, *RAD51D*, *BRIP1*, and *PALB2*.
<sup>□</sup>P-value calculated using Fisher's exact test; Pearson's Chi-squared test; Wilcoxon rank sum test.

### HRD Features, by Self-Reported Race

Germline variants in *BRCA1* were identified in 7% of Black individuals and 10% of White individuals (p=0.45) (Table 1). The majority of germline *BRCA1* variants in Black individuals (7/13) and White individuals (11/12) were nonsense; however, more missense variants were identified in Black individuals (5/13) compared with White individuals (1/12) and a single splice site variant was identified in Black individuals but not White individuals (Figure 1). Almost all germline *BRCA1* variants in White individuals occurred in the large exon 11 (chr17:43094860-43091435, GRCh38) (Figure 2A). In Black individuals, germline *BRCA1* variants were more dispersed and only 4 of 13 variants occurred in exon 11.

**Figure 2.**
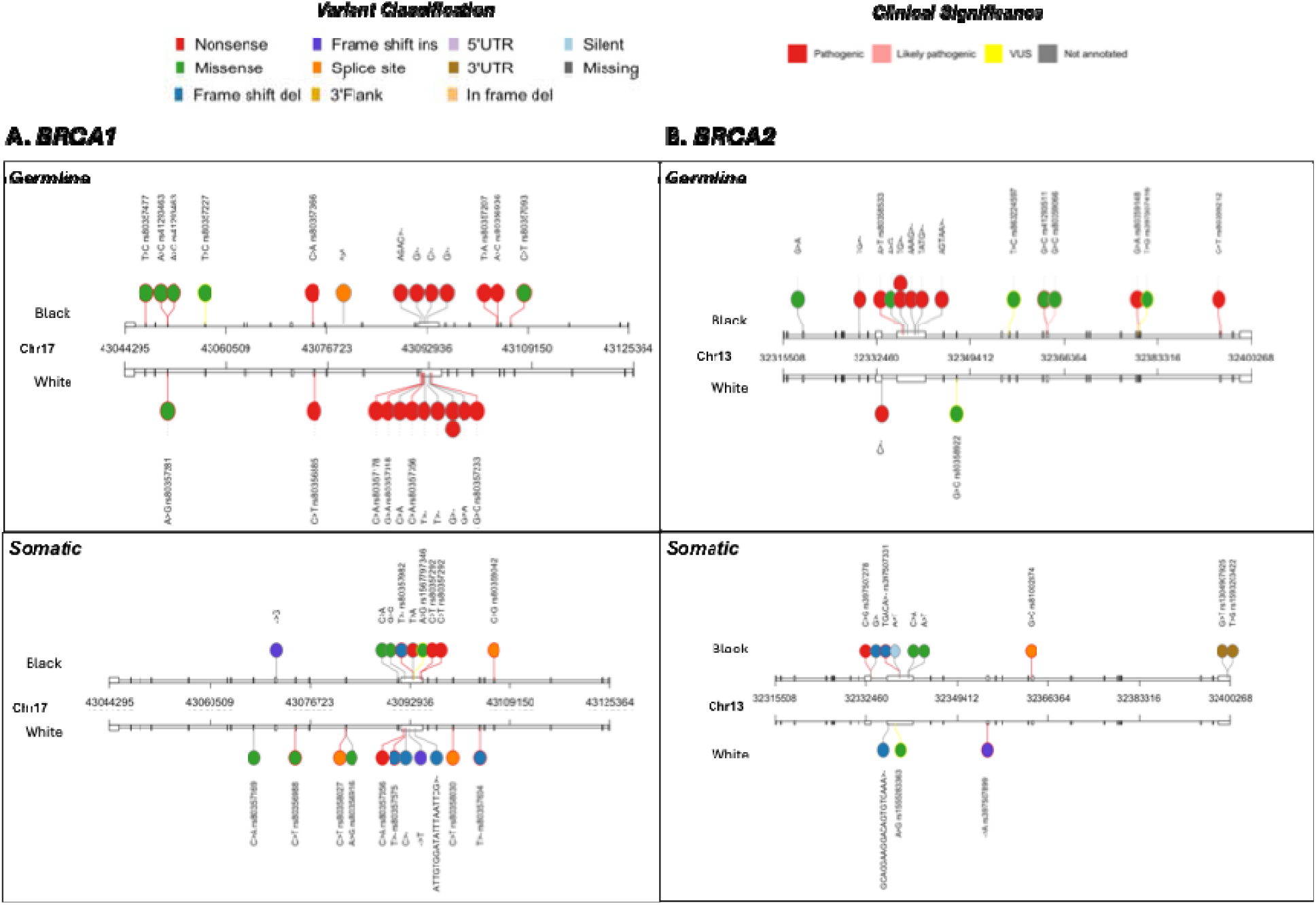
Genomic location, variant classification, clinical significance, and nucleotide change for germline variants and somatic mutations in A) BRCA1 and B) BRCA2 in Black individuals and White individuals. Position defined using GRch38 and exons defined using Transcript ID ENST00000357654 for BRCA1 and ENST00000380152 for BRCA2. Variant classification is indicated by the fill color and clinical significance by the border color for each circle. UTR, untranslated region

The prevalence of germline *BRCA2* variants was higher in Black individuals (8%) compared with White individuals (2%, p=0.012, Table 1, Figure 1). Germline *BRCA2* variants in both groups were missense or nonsense. Over half of the 15 *BRCA2* variants in Black individuals were unannotated, including 6 nonsense and 2 missense variants (Table 2). Other germline *BRCA2* variants in Black individuals included 5 pathogenic or likely pathogenic and 2 VUS both which were missense (Figure 1). Almost half of the germline *BRCA2* variants in Black individuals occurred in the large exon 11 (chr13:32336265-32341196, GRCh38) (Figure 2B). Even after manual review, only two germline variants were identified in White individuals including a missense VUS and an unannotated nonsense variant (Figure 1 and Figure 2B).

**Table 2.**
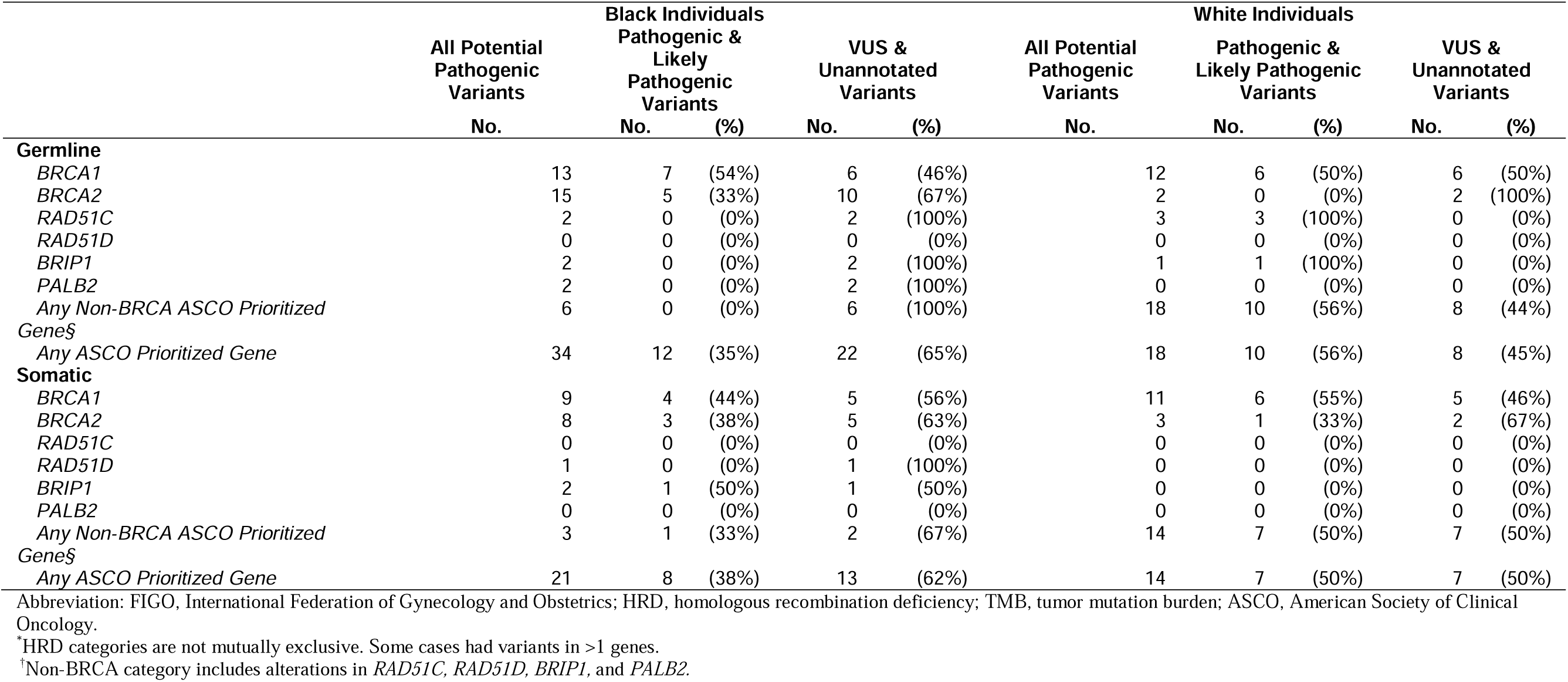
Clinical significance of germline and somatic potential pathogenic variants identified in homologous recombination pathway genes prioritized in ASCO’s recommendations for genetic testing in individuals with HGSC, by self-reported race

|  | All Potential Pathogenic Variants<br>No. | Black Individuals |  | VUS & Unannotated Variants |  | All Potential Pathogenic Variants<br>No. | White Individuals |  | VUS & Unannotated Variants |  |
| --- | --- | --- | --- | --- | --- | --- | --- | --- | --- | --- |
|  |  | Pathogenic & Likely Pathogenic Variants<br>No. | (%) | No. | (%) |  | Pathogenic & Likely Pathogenic Variants<br>No. | (%) | No. | (%) |
| **Germline** |  |  |  |  |  |  |  |  |  |  |
| *BRCA1* | 13 | 7 | (54%) | 6 | (46%) | 12 | 6 | (50%) | 6 | (50%) |
| *BRCA2* | 15 | 5 | (33%) | 10 | (67%) | 2 | 0 | (0%) | 2 | (100%) |
| *RAD51C* | 2 | 0 | (0%) | 2 | (100%) | 3 | 3 | (100%) | 0 | (0%) |
| *RAD51D* | 0 | 0 | (0%) | 0 | (0%) | 0 | 0 | (0%) | 0 | (0%) |
| *BRIP1* | 2 | 0 | (0%) | 2 | (100%) | 1 | 1 | (100%) | 0 | (0%) |
| *PALB2* | 2 | 0 | (0%) | 2 | (100%) | 0 | 0 | (0%) | 0 | (0%) |
| Any Non-BRCA ASCO Prioritized Gene§ | 6 | 0 | (0%) | 6 | (100%) | 18 | 10 | (56%) | 8 | (44%) |
| Any ASCO Prioritized Gene | 34 | 12 | (35%) | 22 | (65%) | 18 | 10 | (56%) | 8 | (45%) |
| **Somatic** |  |  |  |  |  |  |  |  |  |  |
| *BRCA1* | 9 | 4 | (44%) | 5 | (56%) | 11 | 6 | (55%) | 5 | (46%) |
| *BRCA2* | 8 | 3 | (38%) | 5 | (63%) | 3 | 1 | (33%) | 2 | (67%) |
| *RAD51C* | 0 | 0 | (0%) | 0 | (0%) | 0 | 0 | (0%) | 0 | (0%) |
| *RAD51D* | 1 | 0 | (0%) | 1 | (100%) | 0 | 0 | (0%) | 0 | (0%) |
| *BRIP1* | 2 | 1 | (50%) | 1 | (50%) | 0 | 0 | (0%) | 0 | (0%) |
| *PALB2* | 0 | 0 | (0%) | 0 | (0%) | 0 | 0 | (0%) | 0 | (0%) |
| Any Non-BRCA ASCO Prioritized Gene§ | 3 | 1 | (33%) | 2 | (67%) | 14 | 7 | (50%) | 7 | (50%) |
| Any ASCO Prioritized Gene | 21 | 8 | (38%) | 13 | (62%) | 14 | 7 | (50%) | 7 | (50%) |

Abbreviation: FIGO, International Federation of Gynecology and Obstetrics; HRD, homologous recombination deficiency; TMB, tumor mutation burden; ASCO, American Society of Clinical Oncology.
\*HRD categories are not mutually exclusive. Some cases had variants in >1 genes.
†Non-BRCA category includes alterations in *RAD51C*, *RAD51D*, *BRIP1*, and *PALB2*.

Somatic *BRCA1* mutations were identified in 5% of Black individuals and 9% of White individuals (p=0.18). We observed more frame shift deletions in *BRCA1* in White individuals compared with Black individuals (Figure 1 and Figure 2). Somatic *BRCA2* mutations were identified in 5% of Black individuals and 2% of White individuals (p=0.53). The majority of somatic *BRCA2* mutations in Black individuals were unannotated. Unannotated variants included frame shift deletions, missense, and changes in the 3’UTR. A single pathogenic splice site mutation was identified in Black individuals but not White individuals (Figure 1 and Figure 2B). Most somatic *BRCA1* and *BRCA2* mutations occurred in exon 11 of the respective gene in both groups (Figure 2).

The *HRD signature* was observed in 40% of Black individuals compared with 29% of White individuals (p=0.047) (Table 1). Including the HRD signature and a somatic or germline alteration in any ASCO prioritized gene, *any HRD* was observed in 50% of Black individuals and 47% of White individuals (p=0.44).

### HRD Features and tumor mutation burden, age at diagnosis, and survival

Irrespective of self-reported race, germline variants in any *ASCO prioritized gene*, the *HRD signature* and *any HRD* were associated with an increase in TMB (Table 3). Germline and somatic *BRCA1* and *BRCA2* variants were associated with an increase in TMB in Black individuals (Table 3). The median age at diagnosis among individuals with germline *BRCA1* variants was younger than individuals without, and the difference was similar in Black individuals (7.34 years, 95%CI 1.87–12.81) and White individuals (7.76 years, 95%CI 2.30–13.23, Table 3).

**Table 3.** Associations between homologous recombination deficiency features and tumor mutation burden and age at diagnosis, by self-reported race for all potentially pathogenic variants, and restricting to variants of uncertain significance and unannotated variants

| HRD Feature | Median difference between those with HRD feature and those without (95% CI) |  |  |  |  |  |  |  |
| --- | --- | --- | --- | --- | --- | --- | --- | --- |
|  | Median TMB |  |  |  | Age at Diagnosis |  |  |  |
|  | Black (n=178) |  | White (n=123) |  | Black (n=178) |  | White (n=123) |  |
|  | All Potential Pathogenic Variants | VUS & Unannotated Variants | All Potential Pathogenic Variants | VUS & Unannotated Variants | All Potential Pathogenic Variants | VUS & Unannotated Variants | All Potential Pathogenic Variants | VUS & Unannotated Variants |
| **Germline\*** |  |  |  |  |  |  |  |  |
| *BRCA1* |  |  |  |  |  |  |  |  |
| No | Ref | Ref | Ref | Ref | Ref | Ref | Ref | Ref |
| Yes | 0.73 (0.16, 1.31) | 0.78 (0.07, 1.49) | -0.07 (-0.54, 0.39) | -0.18 (-0.77, 0.42) | -7.34 (-12.81, -1.87) | -2.78 (-10.81, 5.25) | -7.76 (-13.23, -2.30) | -10.90 (-17.85, -3.95) |
| *BRCA2* |  |  |  |  |  |  |  |  |
| No | Ref | Ref | Ref | Ref | Ref | Ref | Ref | Ref |
| Yes | 0.53 (0.07, 0.99) | 0.27 (-0.29, 0.83) | 0.36 (-0.72, 1.45) | 0.36 (-0.72, 1.45) | -0.49 (-5.72, 4.73) | -0.30 (-6.6, 6.00) | -6.02 (-19.21, 7.17) | -6.02 (-19.21, 7.17) |
| Non-*BRCA* |  |  |  |  |  |  |  |  |
| ASCO |  |  |  |  |  |  |  |  |
| Prioritized† |  |  |  |  |  |  |  |  |
| No | Ref | Ref | Ref | Ref | Ref | Ref | Ref | Ref |
| Yes | -0.28 (-0.99, 0.44) | -0.28 (-0.99, 0.44) | 1.51 (0.78, 2.23) | 0.64 (-0.12, 1.41) | -1.92 (-9.95, 6.12) | -1.92 (-9.95, 6.12) | 8.35 (-0.96, 17.66) | 1.11 (-8.32, 10.54) |
| Any ASCO |  |  |  |  |  |  |  |  |
| Prioritized |  |  |  |  |  |  |  |  |
| No | Ref | Ref | Ref | Ref | Ref | Ref | Ref | Ref |
| Yes | 0.48 (0.15, 0.81) | 0.33 (-0.07, 0.73) | 0.37 (-0.01, 0.76) | 0.17 (-0.27, 0.62) | -3.58 (-7.27, 0.12) | -1.07 (-5.57, 3.42) | -4.14 (-8.81, 0.54) | -6.84 (-12.14, -1.54) |
| **Somatic\*** |  |  |  |  |  |  |  |  |
| *BRCA1* |  |  |  |  |  |  |  |  |
| No | Ref | Ref | Ref | Ref | Ref | Ref | Ref | Ref |
| Yes | 1.02 (0.45, 1.59) | 1.26 (0.50, 2.02) | 0.24 (-0.24, 0.72) | 0.01 (-0.69, 0.71) | -1.48 (-8.10, 5.14) | -2.87 (-11.64, 5.91) | -4.46 (-10.27, 1.35) | -1.27 (-9.74, 7.20) |
| *BRCA2* |  |  |  |  |  |  |  |  |
| No | Ref | Ref | Ref | Ref | Ref | Ref | Ref | Ref |
| Yes | 0.53 (-0.09, 1.15) | 0.69 (-0.02, 1.41) | 0.07 (-0.82, 0.96) | -0.29 (-1.38, 0.8) | 2.69 (-4.31, 9.68) | 2.57 (-5.46, 10.60) | 4.01 (-6.82, 14.83) | -0.94 (-14.17, 12.29) |
| Any Non-*BRCA* |  |  |  |  |  |  |  |  |
| ASCO |  |  |  |  |  |  |  |  |
| Prioritized† |  |  |  |  |  |  |  |  |
| No | Ref | Ref | Ref | Ref | Ref | Ref | Ref | Ref |
| Yes | -0.02 (-1.03, 0.98) | 0.37 (-0.85, 1.6) | - | * | -0.19 (-11.46, 11.09) | 3.35 (-10.41, 17.11) | - | * |
| Any ASCO |  |  |  |  |  |  |  |  |
| Prioritized |  |  |  |  |  |  |  |  |
| No | Ref | Ref | Ref | Ref | Ref | Ref | Ref | Ref |
| Yes | 0.63 (0.22, 1.04) | 0.79 (0.29, 1.29) | 0.21 (-0.22, 0.64) | -0.08 (-0.67, 0.51) | 0.38 (-4.32, 5.08) | 0.61 (-5.18, 6.39) | -2.65 (-7.90, 2.59) | -1.21 (-8.43, 6.01) |
| **HRD Signature** |  |  |  |  |  |  |  |  |
| No | Ref | Ref | Ref | Ref | Ref | Ref | Ref | Ref |
| Yes | 1.09 (0.88, 1.3) | - | 0.67 (0.39, 0.95) | - | -2.5 (-5.44, 0.43) | - | -2.44 (-6.09, 1.21) | - |
| **Any HRD** |  |  |  |  |  |  |  |  |
| No | Ref | Ref | Ref | Ref | Ref | Ref | Ref | Ref |
| Yes | 1.00 (0.79, 1.21) | - | 0.56 (0.31, 0.82) | - | -4.16 (-7.00, -1.33) | - | -3.96 (-7.24, -0.69) | - |

Abbreviation: FIGO, International Federation of Gynecology and Obstetrics; HRD, homologous recombination deficiency; TMB, tumor mutation burden; ASCO, American Society of Clinical Oncology. \*HRD categories are not mutually exclusive. Some cases had variants in >1 genes. †Non-*BRCA* category includes alterations in *RAD51C*, *RAD51D*, *BRIP1*, and *PALB2*.

Among Black individuals, *BRCA2* variants were associated with better survival (somatic HR=0.23, 95%CI 0.07–0.76; germline HR=0.48, 95%CI 0.22–1.03, Table 4). Germline *BRCA1* variants were associated with worse survival in Black individuals (HR=2.11, 95%CI 1.14–3.88) but not in White individuals (HR=0.89, 95%CI 0.47–1.70). The presence of the *HRD signature* was associated with better survival in both groups (Black HR=0.80, 95%CI 0.55–1.16; White HR=0.81, 95%CI 0.53–1.25), as was *any HRD* (Black HR=0.80, 95%CI 0.56–1.16; White HR=0.76, 95%CI 0.51–1.13). We did not observe a statistically significant interaction between any HRD feature and race (Table 3).

**Table 4.**
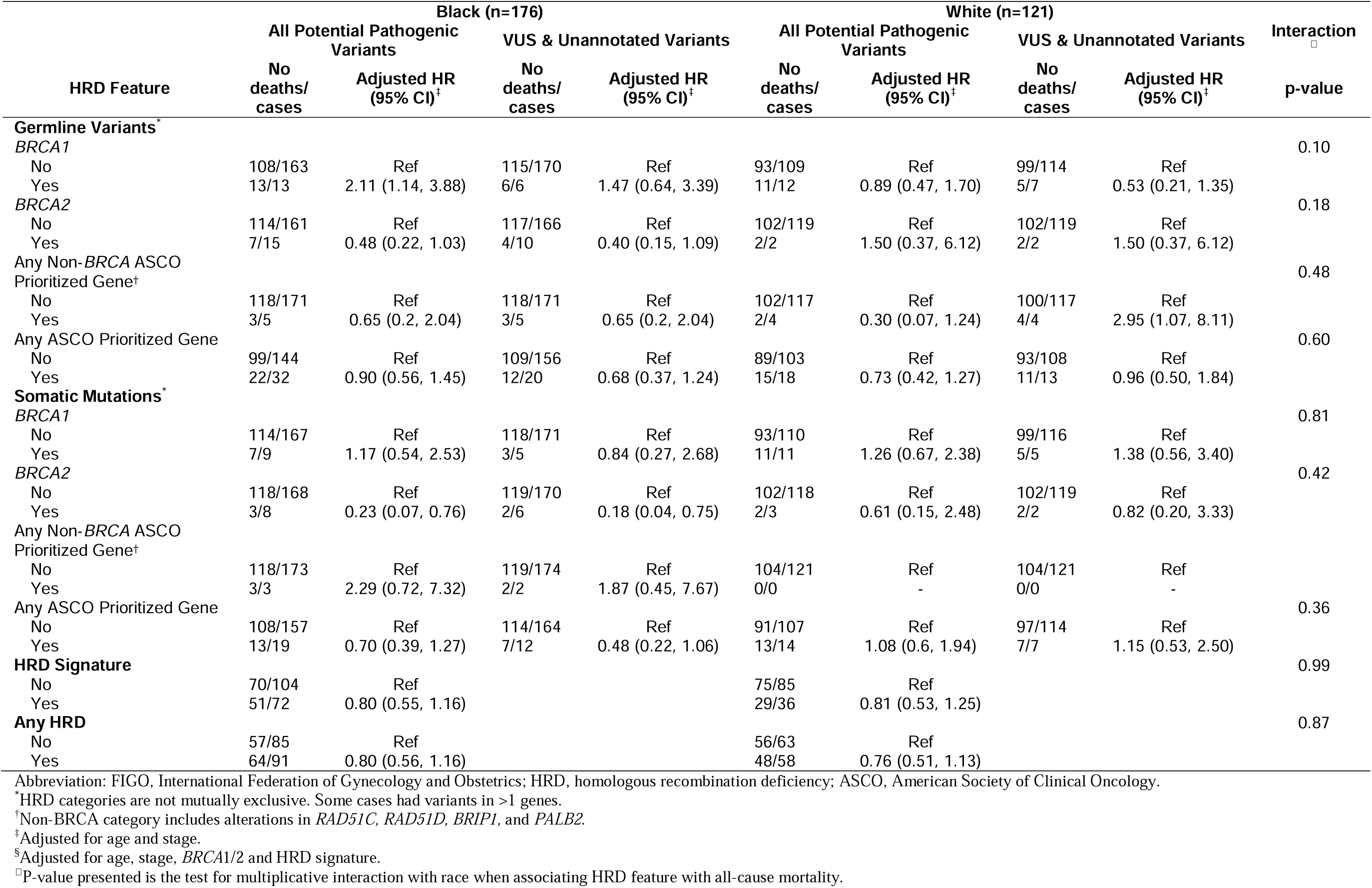
Hazard ratios (HR) and 95% confidence intervals (CI) associating presence of homologous recombination deficiency feature with all-cause mortality, by self-reported race.

### VUS and Unannotated Variants

A higher proportion of germline variants in *any ASCO prioritized gene* were VUS or unannotated in Black individuals (n=22/34, 65%, Figure 1 and Table 2) compared with White individuals (n=8/18, 45%) while a lower proportion were pathogenic or likely pathogenic in Black individuals (n=12/34, 35%) compared with White individuals (10/18, 56%). The proportion of somatic mutations in *any ASCO prioritized gene* that were VUS or unannotated was also higher in Black individuals (n=13/21, 62%) compared with White (n=10/14, 50%).

When we restricted to VUS and unannotated variants, similar patterns were observed for the associations between HRD features and TMB (Table 3). Individuals with germline *BRCA1* variants were diagnosed at an earlier age compared to those without in White individuals (10.9 years, 95%CI 3.95–17.85). *BRCA2* variants continued to be associated with better survival in Black individuals (somatic HR=0.18, 95%CI 0.04–0.75; germline HR=0.40, 95%CI 0.15–1.09, Table 4).

### Other Homologous Recombination Pathway Genes

The prevalence of germline variants in the HRD genes not prioritized in the ASCO recommendations for genetic testing was 11% in Black individuals and 7% in White individuals (p=0.20, eTable 1), and the prevalence of somatic mutations was 11% in Black individuals and 13% in White individuals (p=0.60). Somatic mutations in these genes were associated with an increase in the median TMB in Black individuals (0.54 mutation per Mb, 95%CI 0.06–1.02, eTable 2) and White individuals (0.88 mutation per Mb, 95%CI 0.33–1.44) and were associated with better survival (Black HR=0.52, 95%CI 0.28–0.98; White HR=0.70, 95%CI 0.39–1.25; eTable 3). We did not observe associations between germline variants in these genes and TMB or age at diagnosis. However, germline variants were associated with better survival in Black individuals (HR=0.73, 95%CI 0.39–1.39).

## Discussion

In this study of HRD in HGSC by self-reported race, unselected for family history or mutation status, we found that a greater proportion of variants in HRD genes detected among Black individuals were VUS or unannotated, and that these variants in some HRD genes were associated with survival and differences in the median TMB. While germline and tumor testing for HRD informs precision-based medicine approaches that improve outcomes, a higher proportion of VUS among Black individuals may complicate referral for such care. These findings emphasize the need for continued efforts to characterize variants among racially and ethnically diverse cohorts and to leverage in silico and functional assays to better characterize VUS.

Prior studies have estimated that ∼30% of HGSC have a genomic alteration in homologous recombination pathway genes, most commonly *BRCA1* (germline 9–12% and somatic 3–8%) and *BRCA2* (germline 8% and somatic 3–4%), and another ∼20% have an epigenetic alteration in *BRCA1* or *BRCA2*.^5,39^ In our cohort, the prevalences of any genomic alteration, germline and somatic *BRCA1* variants, and somatic *BRCA2* mutations were similar to prior reports. However, the prevalence of germline *BRCA2* variants was lower in White individuals compared to Black individuals in our study population and compared to prior studies.^5,39^ We also observed that the prevalence of the SBS3 HRD-associated signature was higher in Black individuals compared with White (40% and 29%, respectively). Given that SBS3 does not reflect copy number alterations, it is not unexpected that the prevalence is below 50%, indicating that some individuals were likely misclassified as HRD-negative. However, we would not expect misclassification to be differential, and while differences in sequencing methods, target coverage, and variant filtering may explain differences across studies, these methods were nearly identical for Black and White individuals in our study population. Additionally, it has been reported that there is a higher prevalence of HRD features in breast, lung, and prostate cancers in Black individuals compared to White individuals^30–32^, which is consistent with our observation in HGSC.

Inequities in genetic testing and risk-reducing surgery could have influenced the observed distribution of variants in HRD genes. However, the uptake of genetic testing for *BRCA* among all populations remains relatively low and was higher during the enrollment period for AACES (2010–2015) compared to NCOCS (1999-2005).^40,41^ Additionally, if a higher prevalence of hereditary HGSC was due to lower rates of risk-reducing surgery, we would anticipate observing a higher prevalence of germline variants in *all* genes used in genetic testing. However, germline *BRCA1* variants were more common among White individuals compared with Black individuals. A prior study of families with *BRCA* alterations reported that a higher proportion of *BRCA2* variants among Black individuals localized to genomic regions associated with an elevated ovarian cancer risk compared with variants in White individuals.^20^ This study observed the opposite pattern for *BRCA1* variants with a higher proportion localizing to the elevated risk region in White individuals compared with Black individuals. These findings along with observed variation in *BRCA* variants by geographic region suggest that differences may relate to differences in genetic ancestry.^19–26^

If the prevalence of HRD features differs at a population level, it would be expected that Black individuals would experience better survival. However, racial disparities in survival are well documented and are in part due to inequities in receipt of guideline-concordant care.^42–49^ For example, Black individuals are less likely to receive targeted treatments and be referred for genetic testing due to a family history.^13,14,50^ We observed a higher prevalence of HRD features in Black individuals compared with White, and an identical prevalence of ovarian cancer family history in Black and White individuals, suggesting that at the very least, both populations would benefit equally from genetic testing. However, more variants detected in Black individuals were VUS or unannotated. A higher burden of VUS among Black individuals has been reported previously and is due to differences in genetic variation across populations and inequitable representation in genomics research.^23,24,27–29^ This may diminish the efficacy of genetic testing as VUS are not used for clinical decision-making.^37^ We observed that VUS and unannotated variants were associated with survival suggesting that these variants could be important clinically.

In 2020, ASCO published its guidelines for ovarian cancer germline and tumor genetic testing which included recommending germline testing for *BRCA1, BRCA2*, and other susceptibility genes at the time of diagnosis, and somatic testing for *BRCA1*/*BRCA2* in women without a germline pathogenic or likely pathogenic *BRCA* variant.^37^ Other susceptibility genes prioritized in ASCO recommendation included *RAD51C, RAD51D, BRIP1,* and *PALB2*.^28,51–59^ Other homologous recombination pathway genes not prioritized for genetic testing were mentioned but evidence supporting associations with ovarian cancer risk were inconclusive. Somatic mutations in these genes were associated with higher TMB and better survival among Black individuals highlighting the need to understand these genes in the context of ovarian cancer outcomes.

### Limitations

Individuals in our study were recruited as part of interview-based studies, which likely introduced survivorship bias. However, prior work suggests that survival is similar to SEER populations after conditioning on surviving 10-months, and methods for recruitment, data collection, and genomic assays were nearly identical in Black and White individuals which likely mitigated differential bias.^60^ Whole-exome sequencing may have missed some large *BRCA* rearrangements and the distribution of HRD was not evaluated by genetic ancestry. Sample size limited statistical precision in our analyses and led to imprecise point estimates for some HRD features and survival particularly among White individuals. Further, our study population was limited to Black and White individuals to understand racial disparities in survival for Black individuals, and high rates of VUS have been reported in other racial and ethnic groups. Addressing disparities in understanding of HRD requires further research characterizing HRD across all racial and ethnic groups.

## CONCLUSIONS

Black individuals had a higher prevalence of HRD features associated with better survival, including the HRD signature identified using de novo mutational signature analysis, and germline *BRCA2* variants compared with White individuals. However, more of the germline variants and somatic mutations detected among Black individuals compared with White individuals were VUS or unannotated. These findings suggest that precision medicine strategies for HRD may be particularly important for Black individuals; however, a higher proportion of VUS may complicate referral for such care. Our findings emphasize the need to recruit diverse individuals in genomics research.

## Data Availability Statement

We provide all software under the BSD 3-Clause License. We include scripts for the analyses presented in this manuscript (https://github.com/kmichod/hgsc_hrd_characterization). Deidentified data will be made available through dbGaP under study ID: phs002262.v3.p1.

## Acknowledgments

We would like to acknowledge the AACES interviewers, Christine Bard, LaTonda Briggs, Whitney Franz (North Carolina) and Robin Gold (Detroit). We also acknowledge the individuals responsible for facilitating case ascertainment across the ten sites including: Jennifer Burczyk-Brown (Alabama); Rana Bayakly and Vicki Bennett (Georgia); the Louisiana Tumor Registry; Lisa Paddock and Manisha Narang (New Jersey); Diana Slone, Yingli Wolinsky, Steven Waggoner, Anne Heugel, Nancy Fusco, Kelly Ferguson, Peter Rose, Deb Strater, Taryn Ferber, Donna White, Lynn Borzi, Eric Jenison, Nairmeen Haller, Debbie Thomas, Vivian von Gruenigen, Michele McCarroll, Joyce Neading, John Geisler, Stephanie Smiddy, David Cohn, Michele Vaughan, Luis Vaccarello, Elayna Freese, James Pavelka, Pam Plummer, William Nahhas, Ellen Cato, John Moroney, Mark Wysong, Tonia Combs, Marci Bowling, Brandon Fletcher (Ohio); Martin Whiteside (Tennessee) and Georgina Armstrong and the Texas Registry, Cancer Epidemiology and Surveillance Branch, Department of State Health Services. We would also like to acknowledge the AACES investigators, Anthony J Alberg, Elisa V Bandera, Jill Barnholtz-Sloan, Melissa Bondy, Michele L Cote, Ellen Funkhouser, Edward Peters, Ann G Schwartz, Paul Terry, and Patricia G Moorman. This study would not have been possible without the efforts of the North Carolina Central Tumor Registry and all of the staff of the NCOCS. We also thank Christine Lankevich for her management of the data collection for the North Carolina Study and Rex C. Bentley for review of the pathology in the NCOCS. We would like to thank Rex C. Bentley and Anne M. Mills for the review of the pathology in AACES.

## References

1. Kurman RJ, International Agency for Research on Cancer, World Health Organization. WHO Classification of Tumours of Female Reproductive Organs. vol 6. WHO Classification of Tumours. 2014.

2. Siegel RL, Giaquinto AN, Jemal A. Cancer statistics, 2024. CA Cancer J Clin. Jan-Feb 2024;74(1):12–49. doi:10.3322/caac.21820

3. Peres LC, Cushing-Haugen KL, Kobel M, et al. Invasive Epithelial Ovarian Cancer Survival by Histotype and Disease Stage. J Natl Cancer Inst. Jan 1 2019;111(1):60–68. doi:10.1093/jnci/djy071

4. Lheureux S, Gourley C, Vergote I, Oza AM. Epithelial ovarian cancer. Lancet. Mar 23 2019;393(10177):1240–1253. doi:10.1016/s0140-6736(18)32552-2

5. Cancer Genome Atlas Research N. Integrated genomic analyses of ovarian carcinoma. Nature. Jun 29 2011;474(7353):609–15. doi:10.1038/nature10166

6. Alsop K, Fereday S, Meldrum C, et al. BRCA mutation frequency and patterns of treatment response in BRCA mutation-positive women with ovarian cancer: a report from the Australian Ovarian Cancer Study Group. J Clin Oncol. Jul 20 2012;30(21):2654–63. doi:10.1200/JCO.2011.39.8545

7. Bolton KL, Chenevix-Trench G, Goh C, et al. Association between BRCA1 and BRCA2 mutations and survival in women with invasive epithelial ovarian cancer. Jama. Jan 25 2012;307(4):382–90. doi:10.1001/jama.2012.20

8. Tan DS, Rothermundt C, Thomas K, et al. "BRCAness" syndrome in ovarian cancer: a case-control study describing the clinical features and outcome of patients with epithelial ovarian cancer associated with BRCA1 and BRCA2 mutations. J Clin Oncol. Dec 1 2008;26(34):5530–6. doi:10.1200/jco.2008.16.1703

9. Vencken P, Kriege M, Hoogwerf D, et al. Chemosensitivity and outcome of BRCA1-and BRCA2-associated ovarian cancer patients after first-line chemotherapy compared with sporadic ovarian cancer patients. Ann Oncol. Jun 2011;22(6):1346–1352. doi:10.1093/annonc/mdq628

10. Wilson MK, Pujade-Lauraine E, Aoki D, et al. Fifth Ovarian Cancer Consensus Conference of the Gynecologic Cancer InterGroup: recurrent disease. Ann Oncol. Apr 1 2017;28(4):727–732. doi:10.1093/annonc/mdw663

11. Moore K, Colombo N, Scambia G, et al. Maintenance Olaparib in Patients with Newly Diagnosed Advanced Ovarian Cancer. N Engl J Med. Dec 27 2018;379(26):2495–2505. doi:10.1056/NEJMoa1810858

12. Torre LA, Trabert B, DeSantis CE, et al. Ovarian cancer statistics, 2018. CA Cancer J Clin. Jul 2018;68(4):284–296. doi:10.3322/caac.21456

13. Lin J, Sharaf RN, Saganty R, et al. Achieving universal genetic assessment for women with ovarian cancer: Are we there yet? A systematic review and meta-analysis. Gynecol Oncol. Aug 2021;162(2):506–516. doi:10.1016/j.ygyno.2021.05.011

14. Chapman-Davis E, Zhou ZN, Fields JC, et al. Racial and Ethnic Disparities in Genetic Testing at a Hereditary Breast and Ovarian Cancer Center. J Gen Intern Med. Jan 2021;36(1):35–42. doi:10.1007/s11606-020-06064-x

15. Armstrong K, Micco E, Carney A, Stopfer J, Putt M. Racial differences in the use of BRCA1/2 testing among women with a family history of breast or ovarian cancer. Jama. Apr 13 2005;293(14):1729–36. doi:10.1001/jama.293.14.1729

16. McBride CM, Pathak S, Johnson CE, et al. Psychosocial factors associated with genetic testing status among African American women with ovarian cancer: Results from the African American Cancer Epidemiology Study. Cancer. Mar 15 2022;128(6):1252–1259. doi:10.1002/cncr.34053

17. Cragun D, Weidner A, Lewis C, et al. Racial disparities in BRCA testing and cancer risk management across a population-based sample of young breast cancer survivors. Cancer. Jul 1 2017;123(13):2497–2505. doi:10.1002/cncr.30621

18. Lamacki AJ, Spychalska S, Maga T, et al. Risk-reducing salpingo-oophorectomy among diverse patients with BRCA mutations at an urban public hospital: a mixed methods study. BMJ Open. Jun 17 2024;14(6):e082608. doi:10.1136/bmjopen-2023-082608

19. Vargas E, de Deugd R, Villegas VE, et al. Prevalence of BRCA1 and BRCA2 Germline Mutations in Patients of African Descent with Early-Onset and Familial Colombian Breast Cancer. The Oncologist. 2022;27(2):e151–e157. doi:10.1093/oncolo/oyab026

20. Rebbeck TR, Friebel TM, Friedman E, et al. Mutational spectrum in a worldwide study of 29,700 families with BRCA1 or BRCA2 mutations. Hum Mutat. May 2018;39(5):593–620. doi:10.1002/humu.23406

21. Friebel TM, Andrulis IL, Balmana J, et al. BRCA1 and BRCA2 pathogenic sequence variants in women of African origin or ancestry. Hum Mutat. Oct 2019;40(10):1781–1796. doi:10.1002/humu.23804

22. George SHL, Donenberg T, Alexis C, et al. Gene Sequencing for Pathogenic Variants Among Adults With Breast and Ovarian Cancer in the Caribbean. JAMA Netw Open. Mar 1 2021;4(3):e210307. doi:10.1001/jamanetworkopen.2021.0307

23. Hall MJ, Reid JE, Burbidge LA, et al. BRCA1 and BRCA2 mutations in women of different ethnicities undergoing testing for hereditary breast-ovarian cancer. Cancer. May 15 2009;115(10):2222–33. doi:10.1002/cncr.24200

24. Nanda R, Schumm LP, Cummings S, et al. Genetic Testing in an Ethnically Diverse Cohort of High-Risk WomenA Comparative Analysis of BRCA1 and BRCA2 Mutations in American Families of European and African Ancestry. JAMA. 2005;294(15):1925–1933. doi:10.1001/jama.294.15.1925

25. Sia TY, Maio A, Kemel YM, et al. Germline Pathogenic Variants and Genetic Counseling by Ancestry in Patients With Epithelial Ovarian Cancer. JCO Precis Oncol. Sep 2023;7:e2300137. doi:10.1200/PO.23.00137

26. Wang SM. A global perspective on the ethnic-specific BRCA variation and its implication in clinical application. J Natl Cancer Cent. Mar 2023;3(1):14–20. doi:10.1016/j.jncc.2022.12.001

27. Kurian AW, Ward KC, Abrahamse P, et al. Time Trends in Receipt of Germline Genetic Testing and Results for Women Diagnosed With Breast Cancer or Ovarian Cancer, 2012-2019. J Clin Oncol. May 20 2021;39(15):1631–1640. doi:10.1200/JCO.20.02785

28. Kurian AW, Ward KC, Howlader N, et al. Genetic Testing and Results in a Population-Based Cohort of Breast Cancer Patients and Ovarian Cancer Patients. J Clin Oncol. May 20 2019;37(15):1305–1315. doi:10.1200/jco.18.01854

29. Caswell-Jin JL, Gupta T, Hall E, et al. Racial/ethnic differences in multiple-gene sequencing results for hereditary cancer risk. Genetics in Medicine. 2018/02/01 2018;20(2):234–239. doi:10.1038/gim.2017.96

30. Walens A, Van Alsten SC, Olsson LT, et al. RNA-Based Classification of Homologous Recombination Deficiency in Racially Diverse Patients with Breast Cancer. Cancer Epidemiol Biomarkers Prev. Dec 5 2022;31(12):2136–2147. doi:10.1158/1055-9965.Epi-22-0590

31. Sinha S, Mitchell KA, Zingone A, et al. Higher prevalence of homologous recombination deficiency in tumors from African Americans versus European Americans. Nat Cancer. Jan 2020;1(1):112–121. doi:10.1038/s43018-019-0009-7

32. Petrovics G, Price DK, Lou H, et al. Increased frequency of germline BRCA2 mutations associates with prostate cancer metastasis in a racially diverse patient population. Prostate Cancer Prostatic Dis. Sep 2019;22(3):406–410. doi:10.1038/s41391-018-0114-1

33. Schildkraut JM, Alberg AJ, Bandera EV, et al. A multi-center population-based case-control study of ovarian cancer in African-American women: the African American Cancer Epidemiology Study (AACES). BMC Cancer. Sep 22 2014;14:688. doi:10.1186/1471-2407-14-688

34. Moorman PG, Schildkraut JM, Calingaert B, Halabi S, Vine MF, Berchuck A. Ovulation and ovarian cancer: a comparison of two methods for calculating lifetime ovulatory cycles (United States). Cancer Causes Control. Nov 2002;13(9):807–11. doi:10.1023/a:1020678100977

35. Davidson NR, Barnard ME, Hippen AA, et al. Molecular subtypes of high-grade serous ovarian cancer across racial groups and gene expression platforms. Cancer Epidemiology, Biomarkers & Prevention. 2024;doi:10.1158/1055-9965.Epi-24-0113

36. Lawson-Michod KA MJ, Collin LJ, Nix DA, Davidson N, Huff C, Yao Yu, Atkinson A, Johnson CE, Salas LA, Peres LC, Greene CS, Schildkraut JM, Doherty JA. Characterization of high-grade serous ovarian carcinoma somatic features in Black individuals. In revision with Cancer Research 2024 2024;

37. Konstantinopoulos PA, Norquist B, Lacchetti C, et al. Germline and Somatic Tumor Testing in Epithelial Ovarian Cancer: ASCO Guideline. J Clin Oncol. Apr 10 2020;38(11):1222–1245. doi:10.1200/jco.19.02960

38. Islam SMA, Díaz-Gay M, Wu Y, et al. Uncovering novel mutational signatures by de novo extraction with SigProfilerExtractor. Cell Genom. Nov 9 2022;2(11):None. doi:10.1016/j.xgen.2022.100179

39. Sugino K, Tamura R, Nakaoka H, et al. Germline and somatic mutations of homologous recombination-associated genes in Japanese ovarian cancer patients. Scientific Reports. 2019/11/28 2019;9(1):17808. doi:10.1038/s41598-019-54116-y

40. Knerr S, Bowles EJA, Leppig KA, Buist DSM, Gao H, Wernli KJ. Trends in BRCA Test Utilization in an Integrated Health System, 2005-2015. J Natl Cancer Inst. Aug 1 2019;111(8):795–802. doi:10.1093/jnci/djz008

41. Lau-Min KS, McCarthy AM, Nathanson KL, Domchek SM. Nationwide Trends and Determinants of Germline BRCA1/2 Testing in Patients With Breast and Ovarian Cancer. J Natl Compr Canc Netw. Apr 2023;21(4):351–358.e4. doi:10.6004/jnccn.2022.7257

42. Bristow RE, Powell MA, Al-Hammadi N, et al. Disparities in ovarian cancer care quality and survival according to race and socioeconomic status. J Natl Cancer Inst. Jun 5 2013;105(11):823–32. doi:10.1093/jnci/djt065

43. Bristow RE, Chang J, Ziogas A, Gillen DL, Bai L, Vieira VM. Spatial analysis of advanced-stage ovarian cancer mortality in California. Am J Obstet Gynecol. Jul 2015;213(1):43 e1–43 e8. doi:10.1016/j.ajog.2015.01.045

44. Bristow RE, Chang J, Ziogas A, Campos B, Chavez LR, Anton-Culver H. Impact of National Cancer Institute Comprehensive Cancer Centers on ovarian cancer treatment and survival. J Am Coll Surg. May 2015;220(5):940–50. doi:10.1016/j.jamcollsurg.2015.01.056

45. Bristow RE, Chang J, Ziogas A, Campos B, Chavez LR, Anton-Culver H. Sociodemographic disparities in advanced ovarian cancer survival and adherence to treatment guidelines. Obstet Gynecol. Apr 2015;125(4):833–842. doi:10.1097/AOG.0000000000000643

46. Bristow RE, Chang J, Ziogas A, Anton-Culver H, Vieira VM. Spatial analysis of adherence to treatment guidelines for advanced-stage ovarian cancer and the impact of race and socioeconomic status. Gynecol Oncol. Jul 2014;134(1):60–7. doi:10.1016/j.ygyno.2014.03.561

47. Bristow RE, Chang J, Villanueva C, Ziogas A, Vieira VM. A Risk-Adjusted Model for Ovarian Cancer Care and Disparities in Access to High-Performing Hospitals. Obstet Gynecol. Feb 2020;135(2):328–339. doi:10.1097/AOG.0000000000003665

48. Villanueva C, Chang J, Ziogas A, Bristow RE, Vieira VM. Ovarian cancer in California: Guideline adherence, survival, and the impact of geographic location, 1996-2014. Cancer Epidemiol. Dec 2020;69:101825. doi:10.1016/j.canep.2020.101825

49. Bandera EV, Lee VS, Rodriguez-Rodriguez L, Powell CB, Kushi LH. Racial/Ethnic Disparities in Ovarian Cancer Treatment and Survival. Clin Cancer Res. Dec 1 2016;22(23):5909–5914. doi:10.1158/1078-0432.CCR-16-1119

50. Armstrong DK, Alvarez RD, Bakkum-Gamez JN, et al. Ovarian Cancer, Version 2.2020, NCCN Clinical Practice Guidelines in Oncology. J Natl Compr Canc Netw. Feb 2 2021;19(2):191–226. doi:10.6004/jnccn.2021.0007

51. Song H, Dicks E, Ramus SJ, et al. Contribution of Germline Mutations in the RAD51B, RAD51C, and RAD51D Genes to Ovarian Cancer in the Population. J Clin Oncol. Sep 10 2015;33(26):2901–7. doi:10.1200/jco.2015.61.2408

52. Ramus SJ, Song H, Dicks E, et al. Germline Mutations in the BRIP1, BARD1, PALB2, and NBN Genes in Women With Ovarian Cancer. J Natl Cancer Inst. Nov 2015;107(11) doi:10.1093/jnci/djv214

53. Rafnar T, Gudbjartsson DF, Sulem P, et al. Mutations in BRIP1 confer high risk of ovarian cancer. Nat Genet. Oct 2 2011;43(11):1104–7. doi:10.1038/ng.955

54. Pelttari LM, Heikkinen T, Thompson D, et al. RAD51C is a susceptibility gene for ovarian cancer. Hum Mol Genet. Aug 15 2011;20(16):3278–88. doi:10.1093/hmg/ddr229

55. Norquist BM, Harrell MI, Brady MF, et al. Inherited Mutations in Women With Ovarian Carcinoma. JAMA Oncol. Apr 2016;2(4):482–90. doi:10.1001/jamaoncol.2015.5495

56. Meindl A, Hellebrand H, Wiek C, et al. Germline mutations in breast and ovarian cancer pedigrees establish RAD51C as a human cancer susceptibility gene. Nat Genet. May 2010;42(5):410–4. doi:10.1038/ng.569

57. Loveday C, Turnbull C, Ramsay E, et al. Germline mutations in RAD51D confer susceptibility to ovarian cancer. Nat Genet. Aug 7 2011;43(9):879–882. doi:10.1038/ng.893

58. Lilyquist J, LaDuca H, Polley E, et al. Frequency of mutations in a large series of clinically ascertained ovarian cancer cases tested on multi-gene panels compared to reference controls. Gynecol Oncol. Nov 2017;147(2):375–380. doi:10.1016/j.ygyno.2017.08.030

59. Kanchi KL, Johnson KJ, Lu C, et al. Integrated analysis of germline and somatic variants in ovarian cancer. Nat Commun. 2014;5:3156. doi:10.1038/ncomms4156

60. Schildkraut JM, Johnson C, Dempsey LF, et al. Survival of epithelial ovarian cancer in Black women: a society to cell approach in the African American cancer epidemiology study (AACES). Cancer Causes Control. Mar 2023;34(3):251–265. doi:10.1007/s10552-022-01660-0

