## supplemental materials for "Homologous recombination deficiency in ovarian high-grade serous carcinoma by self-reported race"

### **eTable 1.** Frequencies of germline variants and somatic mutations in HRD genes described in the ASCO literature review, but not prioritized in the ASCO recommendations for genetic testing, by self-reported race

|  | **Black (n=178)** | | **White (n=123)** | |  |
| --- | --- | --- | --- | --- | --- |
|  | **n (%)** | | **n (%)** | | **p-value**^†^ |
| **Germline Variants^*^** | | | | | |
| *ATM* | 5 | (2.8%) | 6 | (4.9%) | 0.4 |
| *MRE11A* | 4 | (2.2%) | 0 | (0%) | 0.15 |
| *RAD50* | 4 | (2.2%) | 0 | (0%) | 0.15 |
| *FANCI* | 3 | (1.7%) | 0 | (0%) | 0.3 |
| *CHEK2* | 2 | (1.1%) | 2 | (1.6%) | >0.9 |
| *RAD51L3* | 1 | (0.6%) | 0 | (0%) | >0.9 |
| *NBN* | 1 | (0.6%) | 0 | (0%) | >0.9 |
| *FANCM* | 1 | (0.6%) | 0 | (0%) | >0.9 |
| Any Gene | 19 | (11%) | 8 | (7%) | 0.2 |
| **Somatic Mutations^*^** | | | | | |
| *ATM* | 1 | (0.6%) | 2 | (1.6%) | 0.6 |
| *ATR* | 2 | (1.1%) | 1 | (0.8%) | >0.9 |
| *ATRX* | 2 | (1.1%) | 3 | (2.4%) | 0.4 |
| *BLM* | 4 | (2.2%) | 1 | (0.8%) | 0.7 |
| *EMSY* | 1 | (0.6%) | 1 | (0.8%) | >0.9 |
| *FANCC* | 1 | (0.6%) | 0 | (0%) | >0.9 |
| *FANCD2* | 1 | (0.6%) | 2 | (1.6%) | 0.6 |
| *FANCE* | 1 | (0.6%) | 0 | (0%) | >0.9 |
| *FANCG* | 1 | (0.6%) | 0 | (0%) | >0.9 |
| *FANCI* | 1 | (0.6%) | 1 | (0.8%) | >0.9 |
| *FANCL* | 1 | (0.6%) | 0 | (0%) | >0.9 |
| *FANCM* | 0 | (0%) | 3 | (2.4%) | 0.07 |
| *RAD50* | 3 | (1.7%) | 0 | (0%) | 0.3 |
| *RAD51* | 1 | (0.6%) | 0 | (0%) | >0.9 |
| *RAD51B* | 0 | (0%) | 1 | (0.8%) | 0.4 |
| *RAD52* | 1 | (0.6%) | 1 | (0.8%) | >0.9 |
| *RAD54L* | 2 | (1.1%) | 0 | (0%) | 0.5 |
| *CHEK2* | 0 | (0%) | 1 | (0.8%) | 0.4 |
| Any Gene | 20 | (11%) | 16 | (13%) | 0.6 |

Abbreviation: FIGO, International Federation of Gynecology and

Obstetrics; HRD, homologous recombination deficiency; ASCO,

American Society of Clinical Oncology.

**^*^**HRD categories are not mutually exclusive. Some cases had variants

in >1 genes.

^†^Pearson's Chi-squared test; Fisher's exact test.

Only genes with a variant or mutation detected are included in the

table. All additional homologous recombination pathway genes

evaluated for germline variants or somatic mutations: *ATM, ATR,*

*ATRX, BARD1, BLM, CHEK1, CHEK2, FANCC, FANCD2, FANCE,*

*FANCF, FANCG, FANCI, FANCL, FANCM, MRE11A, NBN, PALB2,*

*RAD50, RAD51, RAD51B, RAD52, RAD54L, RPA1*.

### **eTable 2**. Associations between HRD features and tumor mutation burden and age at diagnosis by self-reported race, including genes described in the ASCO literature review, but not prioritized in the ASCO recommendations for genetic testing

|  | **Median TMB** | | **Age at diagnosis** | |
| --- | --- | --- | --- | --- |
|  | **Black** | **White** | **Black** | **White** |
|  | **Median difference (95%CI)** | **Median difference (95%CI)** | **Median difference (95%CI)** | **Median difference (95%CI)** |
| **Germline Variants** |  |  |  |  |
| **Any Gene﹡** |  |  |  |  |
| No | Ref | Ref | Ref | Ref |
| Yes | 0.52 (0.24, 0.80) | 0.24 (-0.10, 0.59) | -1.77 (-5.03, 1.49) | -3.89 (-8.05, 0.27) |
| **Somatic Mutations** |  |  |  |  |
| **Any Gene﹡** |  |  |  |  |
| No | Ref | Ref | Ref | Ref |
| Yes | 0.52 (0.20, 0.83) | 0.24 (-0.09, 0.57) | 1.19 (-2.46, 4.84) | -2.30 (-6.32, 1.72) |

Abbreviation: FIGO, International Federation of Gynecology and Obstetrics; HRD, homologous recombination deficiency; ASCO, American Society of Clinical Oncology.

^﹡^﹡Any gene category includes germline variants in *ATM, MRE11A, RAD50, FANCI, CHEK2, RAD51L3, NBN, FANC* and somatic mutations in *ATM, ATR, ATRX, BLM, EMSY, FANCC, FANCD2, FANCE, FANCG, FANCI, FANCL, FANCM, RAD50, RAD51, RAD51B, RAD52, RAD54L, CHEK2*.

### **eTable 3.** Hazard ratio (HR) and 95% confidence interval (CI) associating presence (verses absence) of HRD features with all-cause mortality, for genes described in the ASCO literature review, but not prioritized in the ASCO recommendations for genetic testing

|  | **Black** | | **White** | |
| --- | --- | --- | --- | --- |
|  | **No deaths/cases** | **Adjusted HR (95% CI)**^‡^ | **No deaths/cases** | **Adjusted HR (95% CI)**^‡^ |
| **Germline** |  |  |  |  |
| **Any Gene**﹡ |  |  |  |  |
| No | 110/158 | Ref | 98/114 | Ref |
| Yes | 11/18 | 0.73 (0.39, 1.39) | 6/7 | 0.81 (0.36, 1.86) |
| **Somatic** |  |  |  |  |
| **Any Gene**﹡ |  |  |  |  |
| No | 110/156 | Ref | 91/105 | Ref |
| Yes | 11/20 | 0.52 (0.28, 0.98) | 13/16 | 0.70 (0.39, 1.25) |

Abbreviation: FIGO, International Federation of Gynecology and Obstetrics; HRD, homologous recombination deficiency; ASCO, American Society of Clinical Oncology.

^﹡^﹡Any gene category includes germline variants in *ATM, MRE11A, RAD50, FANCI, CHEK2, RAD51L3, NBN, FANC* and somatic mutations in *ATM, ATR, ATRX, BLM, EMSY, FANCC, FANCD2, FANCE, FANCG, FANCI, FANCL, FANCM, RAD50, RAD51, RAD51B, RAD52, RAD54L, CHEK2*.

^‡^Adjusted for age and stage.

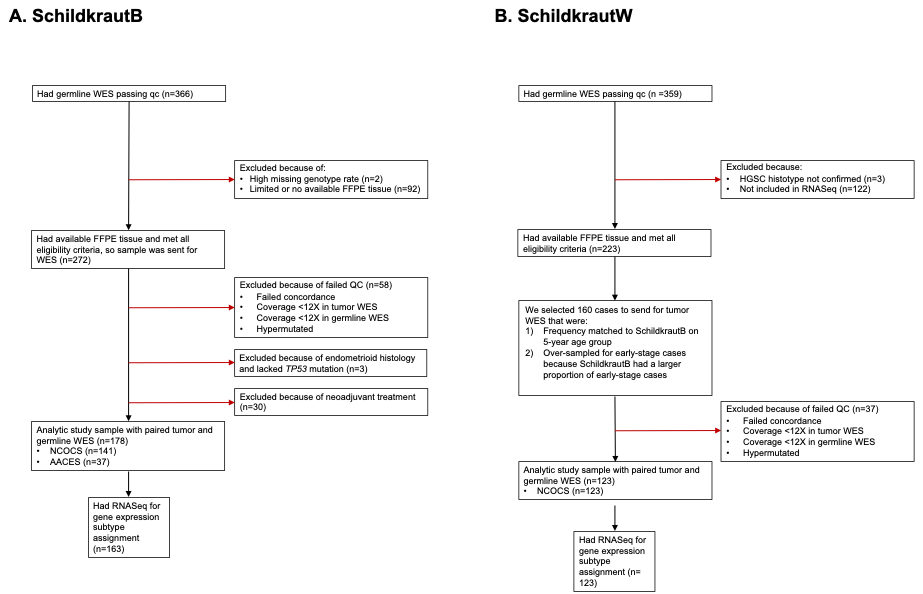

### **eFigure 1.** Flow chart showing inclusion and exclusion criteria for individuals in the A) SchildkrautB and B) SchildkrautW study populations
